# Pediatric solid pseudopapillary neoplasm of the pancreas

**DOI:** 10.1101/2023.07.27.23293297

**Authors:** Ashish Sam Samuel, Deepthi Boddu, Patricia Sebastian, Alex Thomas, T Sreekanth K, Priyanka Hegde, Susan Jehangir

## Abstract

**Background:** Solid pseudopapillary neoplasm of the pancreas (SPN) in children is rare tumor with low malignant potential. Some tumors however behave aggressively. There is very little literature on the management of these variants especially in children. We share our experience of managing large and recurrent SPN and explore the clinicopathological findings correlating to the risk of recurrence.

**Methods:** This is a retrospective study of children who were treated for SPN between 2012 and 2022 at a tertiary care center in India. The clinicopathological features and management strategies in these children were evaluated.

**Results:** 16 children with SPN were treated during this period (88% girls). The median age of presentation was 12 years (IQR 9-14). All children presented with abdominal pain. Computed tomography gave a definitive diagnosis in 81% of cases. The tumor predominantly involved the head of the pancreas (n=9, 56%). Eight of nine children classified as high-grade malignant had a benign course. One child had a recurrence of the tumor 4 years after the initial resection and further recurrence on chemotherapy. She required radiation therapy in addition to reoperation following which she is disease free for 77 months. The overall median follow-up was 46 months (IQR 18-72 months).

**Conclusion:** Complete resection of the tumor provides a cure in most patients with SPN. Recurrent tumors require a multi-modality approach. Long-term survival is good. Better prognostic criteria with immunohistochemistry are required to predict the behavior of these tumors as the WHO criteria for malignancy correlate poorly with clinical outcomes in childhood SPN.

## Introduction

Solid pseudopapillary neoplasm of the pancreas (SPN) is a rare pancreatic tumor that primarily affects young women aged 18 to 35 years, with a female-to-male ratio of 7 - 11:1.(1,2) Approximately 8 to 12% of pediatric pancreatic neoplasms are SPN (1). It is a low malignant tumor with a good prognosis and a 95% long-term disease-free survival rate; however, some cases can be locally aggressive and infiltrative, with metastases to the liver, lung, mesentery, omentum, and/or peritoneum.(3) They lack a specific line of pancreatic epithelial differentiation.^4^ They can undergo a high-grade transformation, characterized by marked nuclear atypia, increased mitoses, and sometimes sarcomatoid foci.(4)

Patients are largely asymptomatic and thus present late, often with large locally infiltrating tumors. Since the first report by Frantz in 1959 several cases have been reported.(5) Herein, we present our experience of 16 children treated for SPN to highlight management challenges for large tumors and recurrent, metastatic variants of this essentially low-grade malignant tumor.

## Methods

The children who were treated for SPN between 2012 and 2022 at the Department of Pediatric Surgery, in a tertiary care center in South India, were included in this retrospective descriptive study. Following approval of the institutional review board (IRB no. 14321), children with SPN were identified by an electronic search of department database and operation records. Computerized medical records were examined for age, gender, clinical presentation, imaging, management, complications, and follow-up.

Each SPN was classified as either low grade malignant neoplasm (LG) or high grade transformation (HG) based on the WHO criteria.(6,7)

Patients were postoperatively classified using the American Joint Committee on Cancer and the Union Internationale Contre le Cancer (AJCC/UICC) TNM residual tumor classification system as R0 (no residual tumor), R1 (microscopic residual tumor), or R2 (macroscopic residual tumor). (8,7,1)

### Statistical Analysis

Frequencies and percentages were calculated for categorical variables and means and standard deviations were calculated for continuous variables. Statistical analysis was done using SPSS v 16 software (Statistical Package for the Social Sciences) using nonparametric analysis.

## Results

Sixteen children with SPN were treated during the study period. Fourteen (88%) were girls. The median age at presentation was 12 years. (IQR 9-14 years). All children sought medical help for abdominal pain. The median size of the tumor at presentation was 6.2cm (IQR 3-8.5). The most common site was the head of pancreas (n=9,56%). Clinical findings classified by site of involvement are summarized in Table 1.

**Table 1.** clinical characteristics based on site of tumor.

| **Characteristic** | **Head\*(n=9)** | **Body (n=3)** | **Tail (n=4)** |
| --- | --- | --- | --- |
| **Age (median)** | **10** | **12** | **13** |
| **Gender** | **m-0, f-9** | **m-1, f-2** | **m-1, f-3** |
| **Presenting complaints** |  |  |  |
| Abdomen pain | 9 | 2 | 4 |
| Vomiting | 6 | 1 | 3 |
| Palpable mass | 1 | 1 | 2 |
| Jaundice | 2 | 0 | 0 |
| Incidental | 0 | 1 | 0 |
| **Size (mean ±SD)** | **5.9+/-2.6** | **5.5+/-2.6** | **7.6+/-2.6** |
| **Imaging findings n(%)** |  |  |  |
| Solid | 1(11) | 1(33) | 0 |
| Cystic | 0 | 1(33) | 0 |
| Mixed | 8(88.9) | 1(33) | 4(100) |

\*including uncinate process

A preliminary Ultrasonogram (US) (Fig. 1) followed by a contrast enhanced Computed tomogram (CT) with oral and IV contrast (Fig. 2) was the chosen mode of evaluation in all children. Selective limited Magnetic Resonance Imaging (MRI) (Fig. 3) was required in eight children to better delineate pancreatic duct anatomy. A definite diagnosis of SPN was made by preoperative imaging in 13 (81%) children; 2 of 9 (22%) on US and 13 of 16 (81%) on CT. Two (13%) tumors were solid, one (6%) cystic and thirteen (81%) mixed solid-cystic architecture. Preoperative biopsy was done in 3 children of whom two children had ultrasound guided biopsy and the other child underwent endoscopic ultrasound guided biopsy.

**Figure 1.**
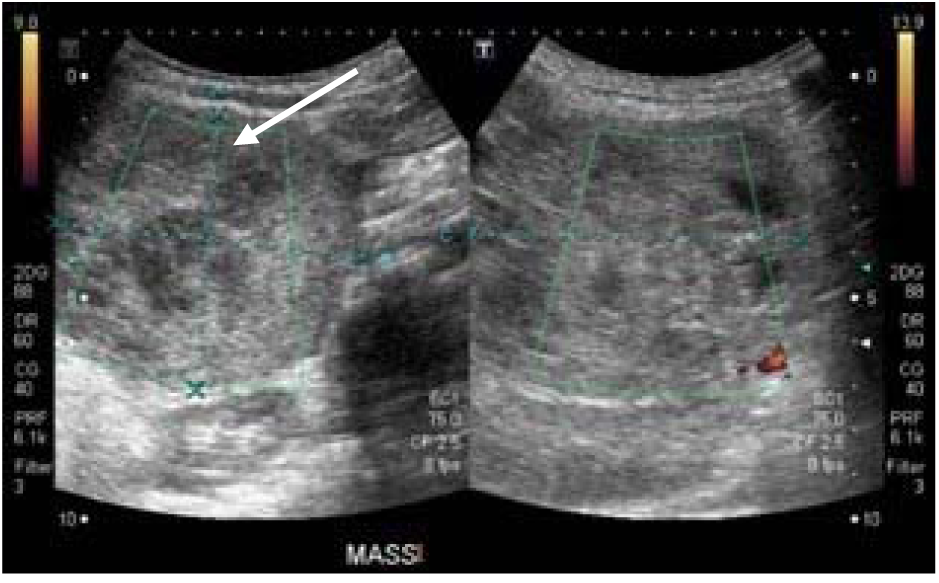
demonstrates ultrasound imaging of typical SPN with arrow pointing cystic area

**Figure 2.**
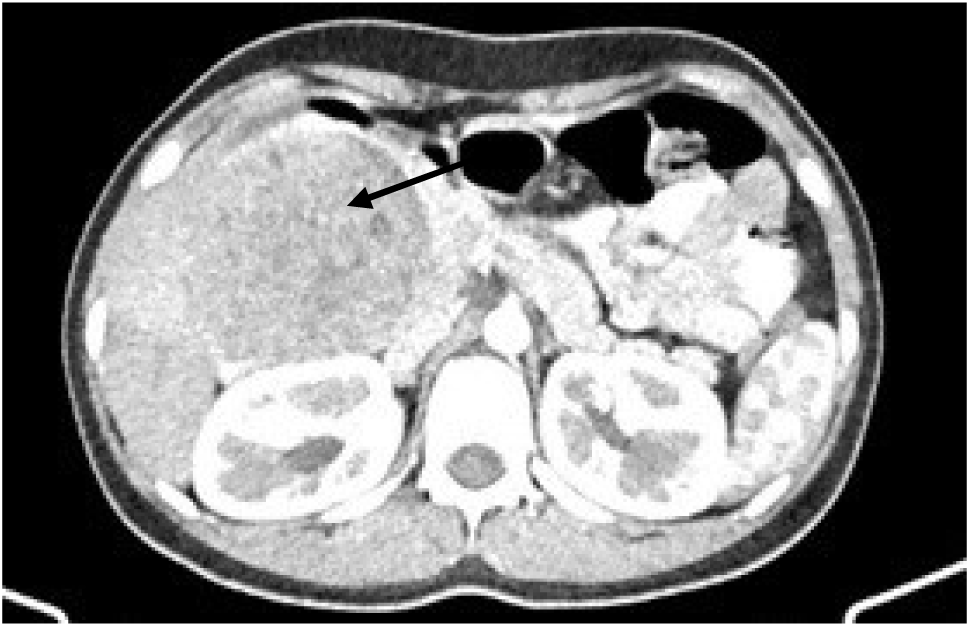
demonstrates a large tumor in the head of the pancreas as seen on computed tomography with arrow pointing at the tumor.

**Figure 3.**
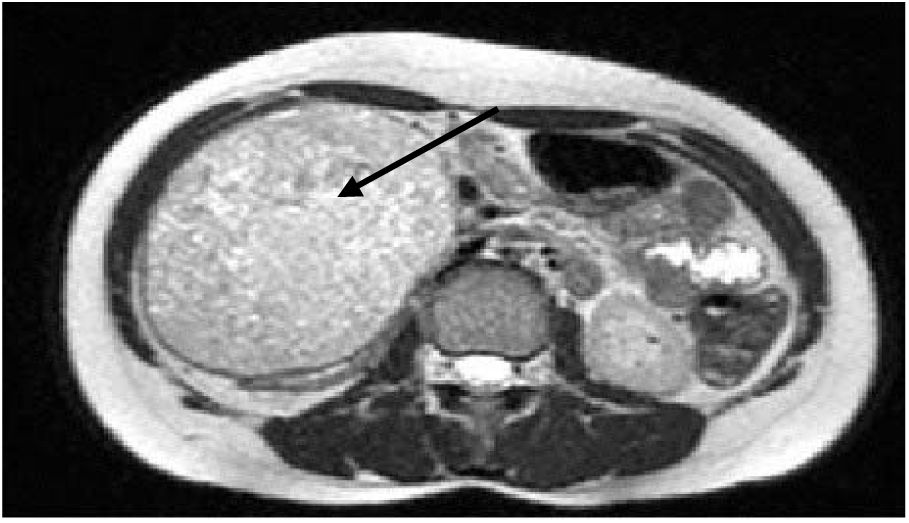
demonstrates a large tumor in the head of the pancreas on MRI with arrow pointing at the tumor.

Of the 9 children with tumor in the head of pancreas, 6 underwent Whipple’s procedure (5 were pylorus preserving) and 3 underwent enucleation of which one was laparoscopic enucleation. Central pancreatectomy was done in one child with tumor in the body of pancreas and rest had enucleation. Distal pancreatectomy was done in 3 children with pancreatic tail involvement.

One child presented with a recurrent abdominal pain and mass 4 years after distal pancreatectomy for SPN at another hospital. CT abdomen, revealed the presence of a mass in the region of pancreatic tail infiltrating the left kidney, spleen, left colon, and a pelvic deposit. She underwent an en-bloc resection of the left kidney, spleen, left colon, remnant tail of pancreas and excision of the pelvic deposit. Following surgery, she was given 2 cycles of chemotherapy with cisplatin and gemcitabine. Repeat imaging at end of two cycles however showed pelvic recurrence. She underwent pelvic metastasectomy, followed by 53 fractions of whole abdomen, intensity modulated radiotherapy (IMRT). She continues to be disease free 6.4 years after completion of treatment.

The other children had localized disease at presentation. The clincopathological findings correlating to the risk of recurrence are enlisted in Table 2. The average maximum diameter of the tumor was 6.25 cm (range 2.2 to 10.5cm) on gross pathology. Six tumors were larger than 8 cm. Resected specimen is shown in (Fig. 4). Three children showed microscopic tumor at the resection margin (R1), one of which was a recurrent tumor. One tumor showed perineural invasion, one had lympho-vascular invasion (recurrence) and nine tumors showed pancreatic parenchymal invasion. The Ki-67 index was available for all 16 patients (Fig. 5) and was >4% (range 4 to 10) in 8 of 16 patients. The child with the recurrent tumor had a Ki-67 of 5-6%. Commonly used immune-histo-chemistry (IHC) markers are the following: CD 10, CD 56, Beta catenin, Cyclin D1, CD 99, Cytokeratin, Chromogranin A, Synaptophysin, Progesterone receptors. In our study all tumors in whom Beta catenin was done showed nuclear positivity. A summary of IHC markers is given in Fig. 5 and Fig 6 is a Photomicrograph displaying pseudo-papillary structures of the neoplasm.

**Figure 4.**
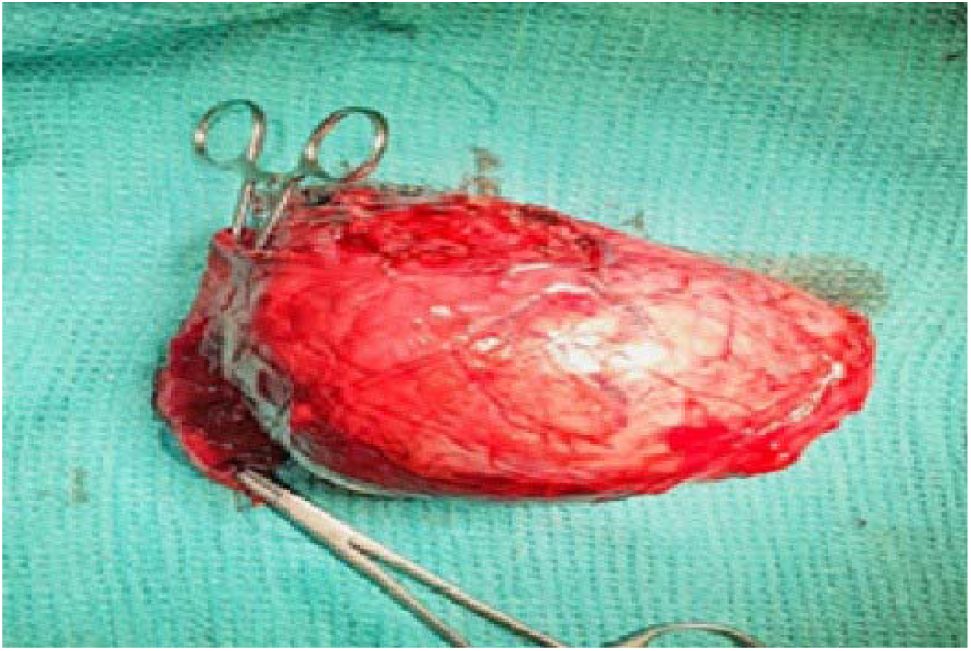
shows the resected tumor with 3rd part of duodenum. The forceps are passed through the duodenum.

**Figure 5.**
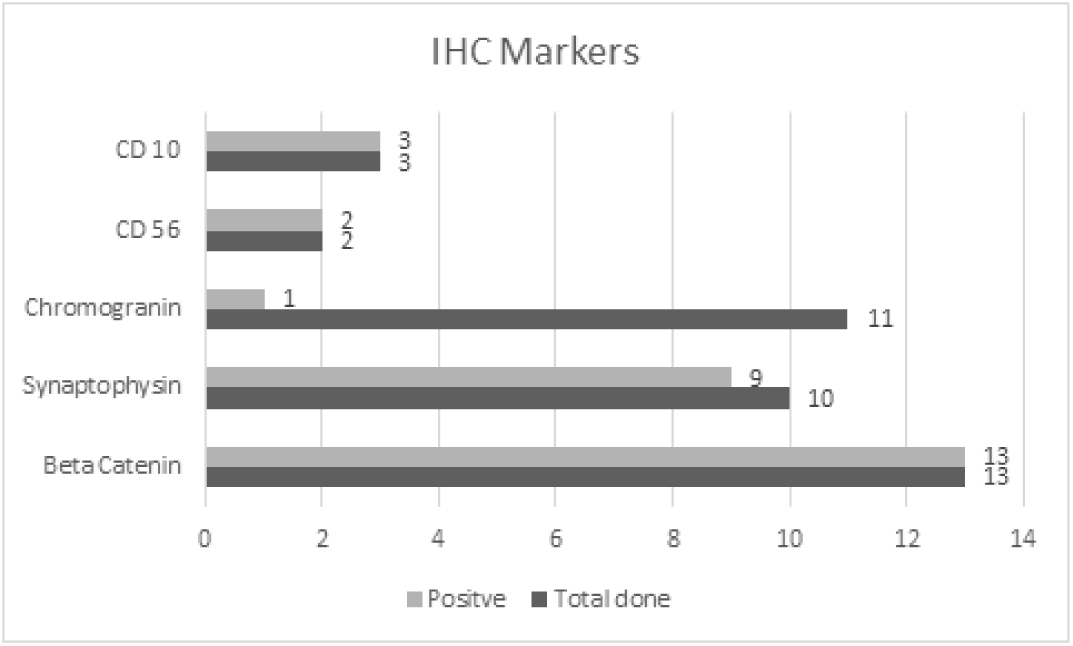
is a graph demonstrating the summary of IHC markers.

**Figure 6.**
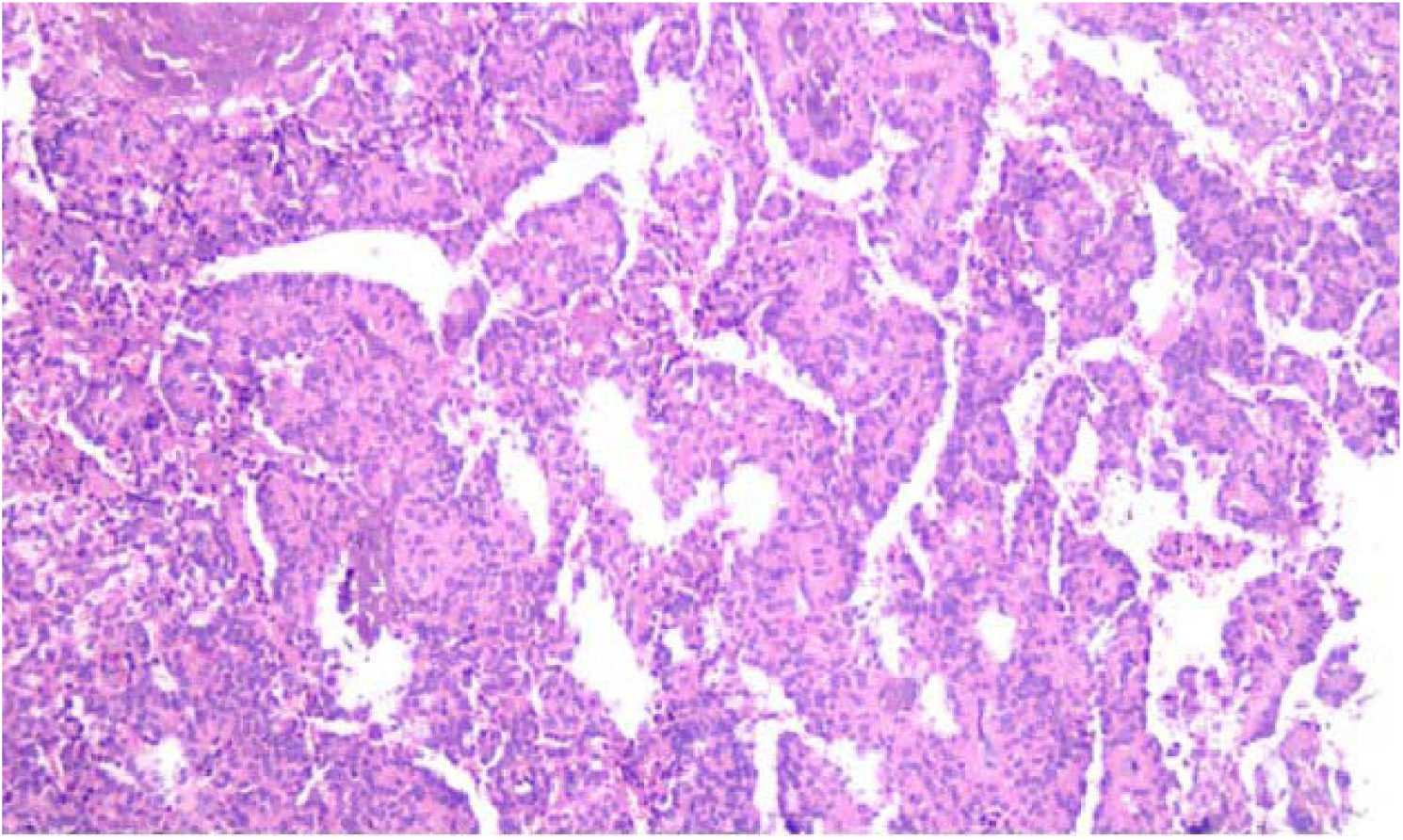
is a Photomicrograph displaying pseudo-papillary structures of the neoplasm, H&E, 10x magnification.

**Table 2.**
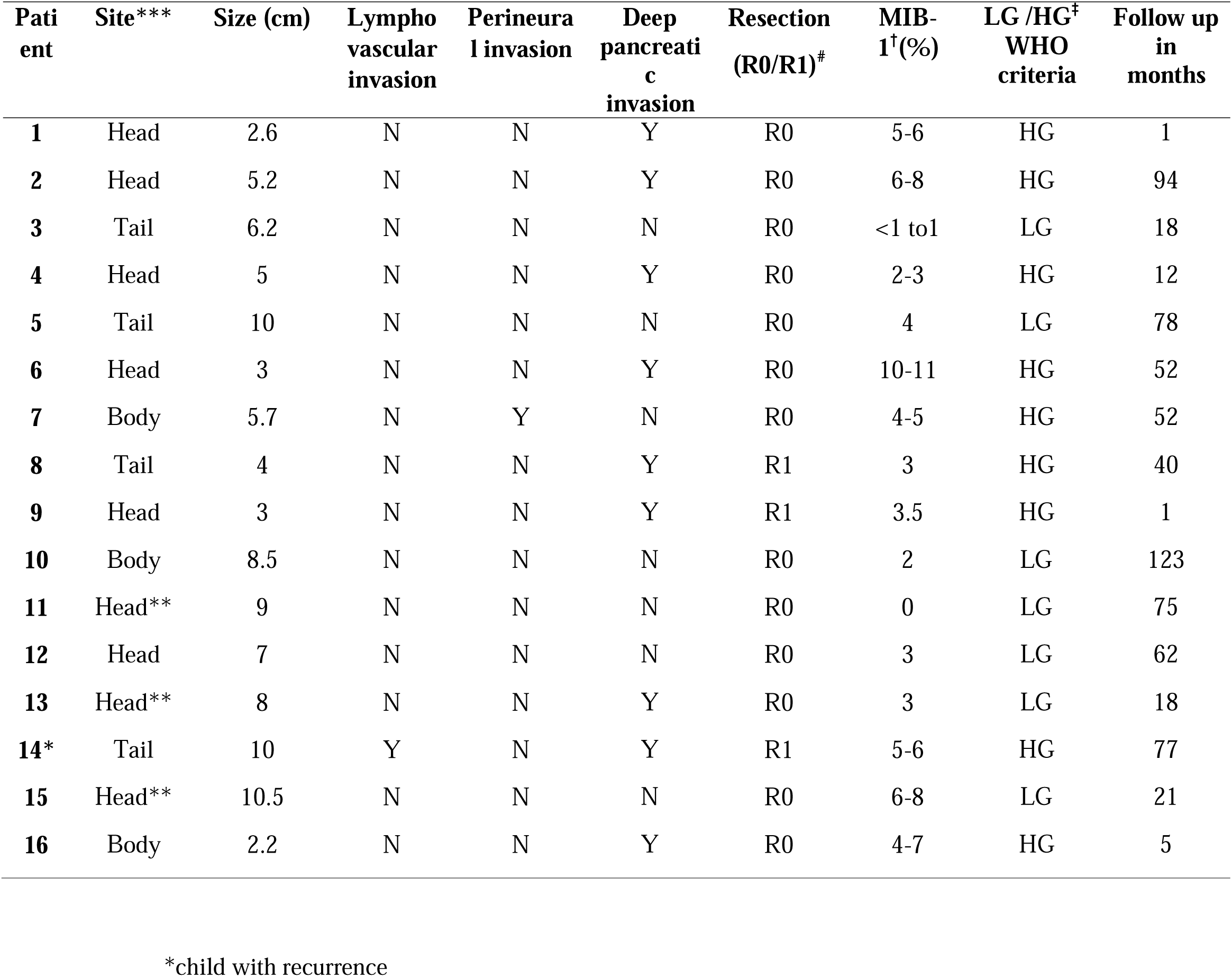
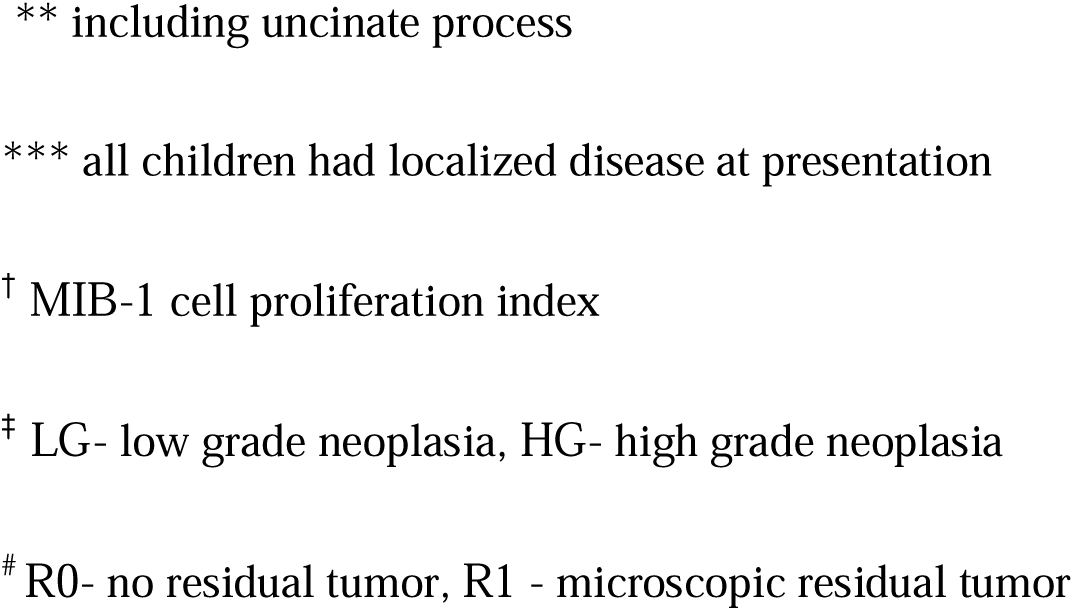
clincopathological findings correlating to the risk of recurrence.

| Patient | Site*** | Size (cm) | Lympho vascular invasion | Perineural invasion | Deep pancreatic invasion | Resection (R0/R1)<sup>#</sup> | MIB-1<sup>†</sup>(%) | LG /HG<sup>‡</sup> WHO criteria | Follow up in months |
| --- | --- | --- | --- | --- | --- | --- | --- | --- | --- |
| 1 | Head | 2.6 | N | N | Y | R0 | 5-6 | HG | 1 |
| 2 | Head | 5.2 | N | N | Y | R0 | 6-8 | HG | 94 |
| 3 | Tail | 6.2 | N | N | N | R0 | <1 to1 | LG | 18 |
| 4 | Head | 5 | N | N | Y | R0 | 2-3 | HG | 12 |
| 5 | Tail | 10 | N | N | N | R0 | 4 | LG | 78 |
| 6 | Head | 3 | N | N | Y | R0 | 10-11 | HG | 52 |
| 7 | Body | 5.7 | N | Y | N | R0 | 4-5 | HG | 52 |
| 8 | Tail | 4 | N | N | Y | R1 | 3 | HG | 40 |
| 9 | Head | 3 | N | N | Y | R1 | 3.5 | HG | 1 |
| 10 | Body | 8.5 | N | N | N | R0 | 2 | LG | 123 |
| 11 | Head** | 9 | N | N | N | R0 | 0 | LG | 75 |
| 12 | Head | 7 | N | N | N | R0 | 3 | LG | 62 |
| 13 | Head** | 8 | N | N | Y | R0 | 3 | LG | 18 |
| 14* | Tail | 10 | Y | N | Y | R1 | 5-6 | HG | 77 |
| 15 | Head** | 10.5 | N | N | N | R0 | 6-8 | LG | 21 |
| 16 | Body | 2.2 | N | N | Y | R0 | 4-7 | HG | 5 |

\*child with recurrence
\*\* including uncinate process
\*\*\* all children had localized disease at presentation
† MIB-1 cell proliferation index
‡ LG- low grade neoplasia, HG- high grade neoplasia

One child developed a pancreatic fistula in the immediate postoperative period. This was managed conservatively and subsided in 18 days with drainage, antibiotics and nutritional support. One child developed a biliary stricture after Whipple’s surgery. She required a reoperation and revision of the biliary enteric anastomosis 1 year after surgery.

The median follow up was 46 months (IQR 18-72 months). 2 patients were lost to follow up after the first postoperative visit at 1 month. On long-term follow-up 3(18.75%) children had pancreatic exocrine insufficiency; none had endocrine insufficiency. One child developed progressive myelopathy after 4 years of surgery. The cause is still under evaluation.

## Discussion

SPN is the most frequent pancreatic tumor in children. It accounts for 1 to3 % of all primary pancreatic tumors.(9) Although it is rare in children, it primarily affects females in their second decade of life.(1,10) Similar to previous studies, our patients were approximately 12 years old, and the male to female ratio was 1:7. (1,2,10,11) Unlike other studies, all children in our group presented with pain rather than being detected incidentally.(4) This finding may be attributed to healthcare-seeking behaviors in developing countries such as India, where caregivers of children typically seek care from informal providers like traditional healers and untrained practitioners first. Only when the issue becomes persistent or worsens do they seek formal medical treatment. (12) Additionally, there is a significant disparity in healthcare facility distribution between urban and rural areas, placing those like our patients at a disadvantage. (12)This could explain the higher incidence of tumors in the head of the pancreas, which are more symptomatic and larger size at presentation than the global trend. SPN, a tumor with unclear etiopathogenesis, has recently garnered attention due to molecular studies. (13,14) The Wnt-CTNNB1 pathway, resulting in the accumulation of β-catenin within the cytoplasm, explains the pathogenesis of at least 90% of SPN cases. Our study found all children with SPN had positive IHC markers for β-catenin. Abnormal functioning of phosphorylating proteins, including GSK3Beta, prevents the degradation of β-catenin, leading to its accumulation in the cytoplasm and eventual migration into the nucleus. In SPN, upregulation of the CTNNB1 gene from the gain of function of the Wnt signaling pathway increases transcription of several target oncogenes, including Cyclin D1, which promotes uncontrolled cell growth and the growth of SPNs. mTOR is the mammalian target for Rapamycin and has been implicated in activating Wnt/Beta catenin signaling pathway.(15,16)

Recent studies have examined the risk factors for recurrence. (17) Recurrence occurs in only 3% of SPN patients following resection, with the majority occurring within five years(3,14,18); consequently, long-term follow-up is essential. Patients with high-risk factors such as, large tumor size (>8cm), synchronous metastasis, lymphovascular invasion, pancreatic parenchymal invasion, and a high Ki-67 index (>4%), are related with recurrence.(10,19–23) The WHO criteria for high grade transformation of SPN include “deep invasion into the surrounding tissue”.(14,24) However, the definition of this criterion may be ambiguous. As a result, we included any pancreatic tissue invasion surrounding the tumor (n=9/16). The child with recurrence in our study had a large tumor with lymphovascular and peripancreatic and pancreatic parenchymal invasion and high Ki 67. However, there was no recurrence in the other 8 patients classified as high grade transformation by the WHO criteria.(21) Except for involvement of an adjacent organ, we believe that pancreatic tissue invasion and capsule invasion may not be associated with local recurrence in children.

The liver has been reported as the most common site of metastatic disease,(25) followed by lymph-nodes and the peritoneum. The median time to recurrence ranged greatly between studies ranging from 1.5 years to 6 years. (21) Improved recurrence prediction for SPN patients could change surveillance strategies, allow for early re-resection, and allow for adjuvant therapy to be considered in high-risk groups of patients. Our patient had a recurrence 4 years after initial surgery, involving the distal pancreas and adjacent spleen, liver and colon and peritoneum metastasis. Close follow-up of this patient could have identified the recurrence early reducing the morbidity of extensive en-bloc resection that was required.

According to the literature, the role of chemotherapy in SPN is two-fold; as neoadjuvant therapy in unresectable tumors (26) and as adjuvant therapy for recurrent or metastatic tumors .(27) There is however, no reliable data on adjuvant or neoadjuvant for this low-grade malignancy. It has been employed in a neoadjuvant role in large tumors (>5cm) in critical locations considered unresectable. Combinations of Cisplatin, 5 fluorouracil and gemcitabine have been used. (26,27) In our series, surgery for the recurrent tumor was followed by two cycles of chemotherapy with Cisplatin and Gemcitabine. However, while on chemotherapy an additional metastatic deposit occurred in the pelvis warranting another operation followed by intensity modulated radiation therapy (IMRT) with a disease-free interval of 77 months. Radiation therapy has been successfully used for recurrent tumors with good long-term prognosis. (28,29) This reiterates that complete to near total excision of tumor in conjunction with other modalities is beneficial in achieving long disease-free period. Drugs that inhibit mTOR and downregulate β-catenin like Sirolimus and Everolimus are showing promise in the treatment of SPN. (16)

Most of the existing information is based on single-institution case series and retrospective research in adults. To verify the prognostic criteria in children and develop the optimum surgical strategy and standard follow-up regimen for SPNs, well-designed multicenter large-scale trials in children with long-term follow-up are required.

## Conclusion

SPN is a rare tumor in children. It manifests clinically in various ways. Although it behaves as benign tumor, some can recur. A complete resection of the tumor provides cure in most cases. If recurrence is noted, further surgical clearance and adjuvant therapy provides good long-term results. Better prognostic criteria with immunohistochemistry are required to predict the behavior of these tumors as the WHO criteria for high grade malignancy correlate poorly with clinical outcome in childhood SPN.

## Data Availability

All data produced in the present study are available upon reasonable request to the authors

## Abbreviations key

|  |  |
| --- | --- |
| SPN | solid pseudopapillary neoplasm |
| LG | Low grade |
| HG | High grade |
| AJCC/UICC | American Joint Committee on Cancer / International Union Against Cancer |
| TNM | Tumor, node, metastasis |
| IQR | Interquartile range |
| US | Ultrasonogram |
| CT | Computed tomogram |
| MRI | Magnetic resonance imaging |
| IV | Intravenous |
| IMRT | Intensity modulated radiation therapy |
| WHO | World health organisation |
| IHC | Immuno histo chemistry |
| MIB-1/Ki-67 | Cell proliferation index |

## Conflict of interest

The authors declare no conflict of interest

## Notes

**Conflict of interest:** Nil

### Competing Interest Statement

The authors have declared no competing interest.

### Funding Statement

This study did not receive any funding

### Author Declarations

Ethics committee/IRB of Christian Medical College, Vellore gave ethical approval for this work

